# The Seasonal Influenza Vaccine Cannot Trigger a Titer Increase Among Some Elderly Individuals

**DOI:** 10.1101/2024.01.17.24301451

**Authors:** Yang Ge, Wangnan Cao, Shengzhi Sun, Ted M. Ross, Ye Shen

## Abstract

Many elderlies exhibited absent responses to influenza vaccines. Our exploration of this heterogeneity revealed associations with vaccine dose (HD vs. SD, OR: 0.59 (95%CrI, 0.4 to 0.87)), pre-vaccination titer levels (OR: 1.57 (95%CrI, 1.38 to 1.8), and gender (Male vs. Female OR: 2.12 (95%CrI, 1.38 to 3.25)).

## Main text

Influenza results in over half a million deaths annually, predominantly among older adults [1,2]. Certain older individuals exhibit a limited or absent response to vaccines, while others do not. Investigating the factors contributing to this disparity can enhance our comprehension of high-risk populations and facilitate the enhancement of vaccination strategies.

According to CDC’s recommendations [3], adults aged 65 years and older have the option to choose a high-dose (HD) influenza vaccine over a standard-dose (SD) one. The licensed SD split-inactivated virion vaccine contains 15 micrograms of HA antigen for each strain. In contrast, Sanofi Pasteur’s Fluzone HD provides 60 micrograms of HA for each vaccine strain, showing a relative benefit [4]. It would be intriguing to explore this benefit further, investigating whether the HD vaccine reduces the proportion of older adults with a limited or absent response to vaccines or merely enhances antibody levels in those who respond.

We obtained human vaccination data covering the flu seasons from 2014/15 to 2019/20 from our ongoing vaccination cohort study [5,6]. Each year, investigators enrolled individuals who hadn’t received an influenza vaccine in the season preceding the flu season’s onset. People aged 65 and older were offered a choice between HD and SD vaccines. Serological samples were collected both before vaccination and 21-28 days after. Hemagglutination-inhibition (HAI) antibody levels were assessed using standard dilution assays. Thus, the data has hierarchical features, with individuals repeatedly enrolled in different flu seasons and tested for HAI titers for different homologous strains. The HD vaccine was trivalent during the entire study years, while the SD vaccines were trivalent in 2013 and 2014, but quadrivalent in later years, with the quadrivalent formulation containing an additional B-strain. Unlike the SD, the trivalent HD vaccine contained only a single B strain; therefore, in this study, we focused solely on influenza A subtypes and excluded results related to all B strains. Our emphasis was on the homologous HAI titer increase of the vaccine. It was defined as log2 scale of the ratio of the post-vaccination Hemagglutination Inhibition (HAI) titer over the pre-vaccination HAI titer. This outcome signifies the difference between post-vaccination and pre-vaccination titers, providing insight into a participant’s immunological response to the vaccine.

The HAI titer, obtained through the dilution assay, exhibited discrete positive values, resulting in discrete observations of titer increase. However, a significant number of participants showed no titer increase after vaccination, leading to an excess of zeros and overdispersion in the outcome of titer increase. To address this, we assumed the elderly could be categorized into two sub-populations. The first, **Sensitive to the Influenza Vaccine (SIV)**, included participants with a positive vaccine-induced titer increase. The other, **Insensitive to the Influenza Vaccine (IIV)**, displayed either zero or negative titer increase. The HAI titer, obtained through the dilution assay, had discrete positive values, leading to discrete observations of titer increase. Thus, we applied a hurdle Poisson regression model to our data.

Our hurdle Poisson regression model combines a logistic and Poisson process. The logistic process generates zero to represent the IIV sub-population, characterized by zero or negative titer increase. When the logistic process yields one (representing the SIV sub-population), the Poisson process is used to model the positive values of titer increase. If we define *θ* as the probability of IIV and *λ* as the titer increase for SIV, the probability mass function of the hurdle-Poisson model is defined by

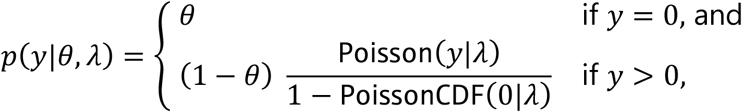

To assess the vaccine dose impact (HD vs. SD) while accounting for multilevel features in the study, we used a Bayesian multilevel modeling approach. This included incorporating two random intercepts for repeated enrollment and strains formulated into a single vaccine. Our models also considered age, gender, race, and pre-vaccination titer as covariates. We use mean and credible interval (CrI) to summarize results.

Our samples cover influenza seasons from 2013/14 to 2019/20. During this time, the HD and SD vaccines were given for 564 and 440 times, respectively, to individuals aged 65 years and older (Figure 1A). Both vaccine groups had comparable age and gender distributions, along with similar median pre-vaccination titers below 1:40. Across the entire study period, eight different influenza type A vaccine strains were observed. About one-third of the elderly population did not show a positive titer increase after vaccination, ranging from 6% to 53% (Figure 1B).

**Figure 1:**
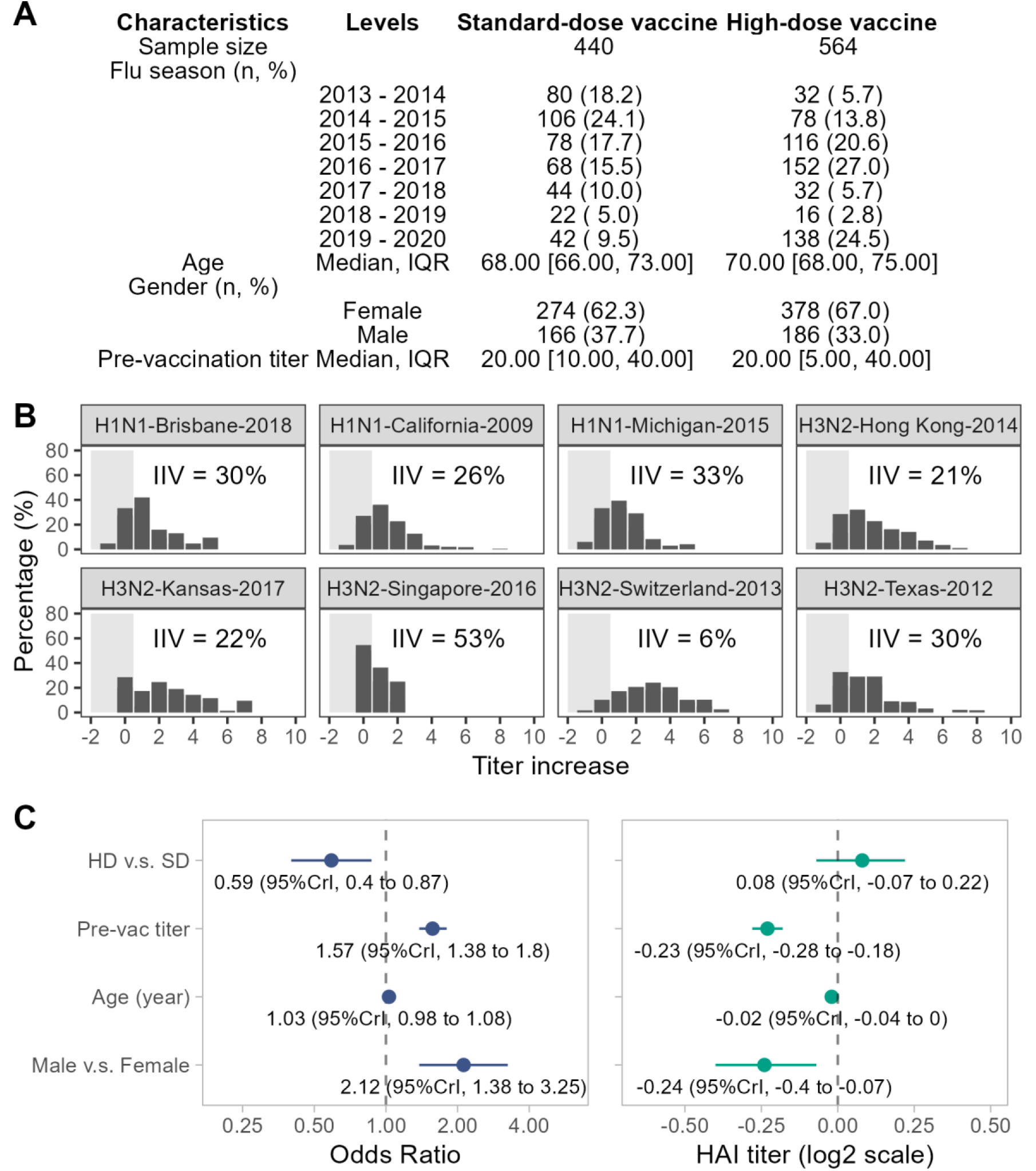
Data description and hurdle-poisson model outputs. A) Data description; B) The distribution of titer increase for different vaccine strains. The proportions of Elderly sub-population were insensitive to the influenza vaccine (IIV) has been calculated. C) Estimates of the hurdle-poisson model. The left side is the logistic model for estimatingthe associations between variables and the likelihood of being an IIV individual. The right side is the Poisson model for estimating the associations between variables and the titer increase for an SIV individual.

The results of the hurdle-Poisson model are shown in Figure 1C. In comparison to the SD, individuals who received the HD vaccine had an Odds Ratio (OR) of being an IIV as 0.59 (95%CrI, 0.4 to 0.87). Additionally, higher pre-vaccination titer and being male had ORs equal to 1.57 (95%CrI, 1.38 to 1.8) and 2.12 (95%CrI, 1.38 to 3.25), respectively. In cases where an individual responded to the influenza vaccine, HD might associate with a small additional titer increase as 0.08 (95%CrI, -0.07 to 0.22). Conversely, higher pre-vaccination titer and being male were associated with a reduced titer increase as -0.23 (95%CrI, -0.28 to -0.18) and -0.24 (95%CrI, -0.4 to -0.07), respectively.

Influenza vaccine effectiveness varies among populations, showing significantly reduced protection (10%) among the elderly [7]. An important reason could be aligned with our findings that many elderly individuals (> 30%) had limited or absent immune responses after vaccination. Our study suggests that the HD vaccine may decrease the likelihood of an elderly individual becoming an IIV, but it has only a mild impact on the actual titer increase if a response is triggered. We also observed that individuals with some pre-existing immunities or being male had a worse response to the vaccine than their counterparts.

Our study controlled for repeated influenza vaccination and the administration of multiple strains simultaneously in trivalent or quadrivalent vaccines. However, our sample size was not balanced across the seven influenza seasons, and participants were not randomly assigned to HD or SD groups, potentially introducing bias. Additionally, we were not able to further explore why some individuals did not adequately respond to the vaccine due to a lack of detailed health background information.

Understanding the mechanism behind how individuals become the IIV population poses a significant challenge, but it is crucial for the development of a universal influenza vaccine [8]. If an influenza vaccine only works on the low-risk population and cannot induce an immune response in high-risk populations (e.g., the elderly), our fight against the impact of the flu remains challenging and arduous. Additional research is warranted to study why the influenza vaccine cannot trigger a titer increase among some elderly individuals.

## Data Availability Statement

The data supporting the findings of this study are available upon reasonable request.

## Acknowledgements

The authors would like to thank Hannah Hanley for coordinating the UGA influenza vaccine study, as well as Brittany Baker, Frankie Stewart, and Emma Whitesell for excellent phlebotomy and data entry services. The authors would also like to thank Hana Ji, James Allen, Michael Carlock, Jasmine Burris, Omar Hamwy, Mitchell Lee, Laruen Howland for processing samples and Rosemary Kim, Jasper Gattiker, and Erin Jarrett for performing and virological assays. The authors also thank Brad Phillips and Kim Schmitz from the UGA Clinical and Translation Research Unit for assistance in enrollment and sample collections. Finally, the authors thank all the enrolled participants for providing their time and samples to the study.

